# Evidence for non-Mendelian inheritance in spastic paraplegia 7

**DOI:** 10.1101/2020.09.25.20176032

**Authors:** Mehrdad A Estiar, Eric Yu, Ikhlass Haj Salem, Jay P. Ross, Kheireddin Mufti, Fulya Akçimen, Etienne Leveille, Dan Spiegelman, Jennifer A. Ruskey, Farnaz Asayesh, Alain Dagher, Grace Yoon, Mark Tarnopolsky, Kym M. Boycott, Nicolas Dupre, Patrick A. Dion, Oksana Suchowersky, Jean-Francois Trempe, Guy A. Rouleau, Ziv Gan-Or

## Abstract

Hereditary spastic paraplegia is a group of rare motor neuron diseases considered to be inherited in a classical monogenic Mendelian manner. Although the typical inheritance of spastic paraplegia type 7 is autosomal recessive, several reports have suggested that *SPG7* variants may also cause autosomal dominant HSP. We aimed to conduct an exome-wide genetic analysis on a large Canadian cohort of hereditary spastic paraplegia patients and controls to examine the association of *SPG7* and hereditary spastic paraplegia. In total, 585 hereditary spastic paraplegia patients from 372 families and 1,175 controls, including 580 unrelated individuals, were analyzed for the presence of *SPG7* variants. Whole exome sequencing was performed on 400 hereditary spastic paraplegia patients (291 index cases) and all 1,175 controls. After excluding 38 biallelic hereditary spastic paraplegia type 7 patients, the frequency of heterozygous pathogenic/likely pathogenic *SPG7* variant carriers (4.8%) among hereditary spastic paraplegia unrelated index cases who underwent WES, was significantly higher than among unrelated controls (1.7%; OR=2.88, 95%CI=1.24-6.66, *p*=0.009). The heterozygous *SPG7* p.(Ala510Val) variant was found in 3.7% of index cases vs. 0.85% in unrelated controls (OR=4.42, 95%CI=1.49-13.07, *p*=0.005). We identified four heterozygous *SPG7* variant carriers with an additional pathogenic variant in genes known to cause hereditary spastic paraplegia, compared to zero in controls (OR=19.58, 95%CI=1.05-365.13, *p*=0.0031; Fisher’s Exact test with Haldane-Anscombe correction), indicating potential digenic inheritance. We further identified four families with heterozygous variants in *SPG7* and SPG7-interacting genes *(CACNA1A, AFG3L2* and *MORC2)*. Out of these, there is especially compelling evidence for epistasis between *SPG7* and *AFG3L2*. The p.(Ile705Thr) variant in AFG3L2 is located at the interface between hexamer subunits, in a hotspot of mutations associated with spinocerebellar ataxia type 28 that affect its proteolytic function. Our results provide evidence for complex inheritance in SPG7-associated hereditary spastic paraplegia, which may include recessive and possibly dominant and digenic/epistasis forms of inheritance.

## Introduction

Hereditary spastic paraplegia (HSP) is a group of rare neurodegenerative diseases considered to be inherited in a classical monogenic Mendelian manner. To date, more than 80 loci or genes have been implicated in HSP, and some of these genes are also known to be involved in diseases whose typical features differ from HSP (Synofzik *et al.*, 2014). Patients diagnosed with HSP may present with a wide spectrum of symptoms, from very subtle lower limb spasticity to severe neurological and non-neurological manifestations. These symptoms often overlap with other disorders, which may lead to incorrect diagnoses (Diomedi *et al.*, 2016; Leveille *et al.*, 2018).

Spastic paraplegia 7 (SPG7, OMIM # 607259) is the first-identified autosomal recessive (AR) type of HSP, accounting for 5-12% of AR-HSP (Brugman *et al.*, 2008). Pathogenic variants in *SPG7* (OMIM # 602783) may also lead to spastic and cerebellar ataxia, and they were also associated with peripheral neuropathy with no other neurological symptoms, primary progressive multiple sclerosis, amyotrophic lateral sclerosis, primary lateral sclerosis, parkinsonism, limb dystonia, and isolated dominant optic atrophy (Klebe *et al.*, 2012; Mitsumoto *et al.*, 2015; Krüger *et al.*, 2016; Schaefer *et al.*, 2018; De la Casa-Fages *et al.*, 2019; Liu *et al.*, 2019; Bellinvia *et al.*, 2020; Charif *et al.*, 2020; Osmanovic *et al.*, 2020). Although the typical inheritance of *SPG7* is considered to be AR, several reports have suggested that *SPG7* variants may also cause autosomal dominant (AD) HSP (McDermott *et al*, 2001; Arnoldi *et al.*, 2008; Klebe *et al.*, 2012; Sanchez-Ferrero *et al.*, 2013). In none of these studies was the possibility of digenic inheritance (carriers of two mutations in 2 distinct genes associated with HSP) examined, therefore the role of *SPG7* in AD-HSP remains controversial and requires additional studies.

*SPG7* encodes the paraplegin/SPG7 protein, localized at the inner mitochondrial membrane. SPG7 is part of the m-AAA metalloproteinase complex that has roles in diverse cellular processes including organelle biogenesis, intracellular motility, membrane trafficking, protein folding, and proteolysis. The *SPG7* gene is comprised of 17 exons with mutational hotspots in exons 11, 13 and 15 (Coarelli *et al.*, 2019). The p.(Ala510Val) variant in exon 11 is a known mutational hotspot in *SPG7*, which despite relatively high allele frequency (AF) in publicly available databases (0.0027 in control samples in gnomAD), is considered pathogenic (Bonn *et al.*, 2010; Klebe *et al.*, 2012; Sánchez Ferrero *et al.*, 2013), likely with incomplete penetrance or variable expressivity. The high frequency of the p.(Ala510Val) variant, the wide clinical spectrum, and the possibility of dominant and recessive transmission make genetic counseling and correct diagnosis challenging in SPG7.

In the current study, we performed a comprehensive genetic analysis in a large cohort of HSP patients with either mono-allelic or bi-allelic variants in *SPG7*. We examined whether heterozygous *SPG7* variants are overrepresented in HSP patients, and whether digenic inheritance of *SPG7* variants together with other HSP-related gene variants may have a role in HSP.

## Materials and methods

### Population

A total of 585 HSP patients from 372 families and 1,175 control individuals have been recruited across Canada and were analyzed for the presence of *SPG7* variants. Initially, 379 HSP patients were analyzed using a sequencing panel of known HSP genes. Out of those, 194 were not genetically diagnosed, and went through whole-exome sequencing (WES). An additional 206 HSP patients and 1,175 individuals who served as a control group (including healthy individuals and patients with non-movement disorders) were sequenced directly using WES. Of the 1,175 controls, 580 were unrelated individuals. Out of all the unrelated control individuals (n=580) and HSP index cases (n=372), 70% and 88.4% were Europeans, respectively, according to the HapMap Project data (Nature, 2007). Principal component analysis (PCA, supplementary figure 1), shows overlapping main principle components. Since we study very rare variants in cases and controls, ethnicity is likely to have no or very minor effect. Clinical characteristics of HSP patients and controls are detailed in Supplementary Tables 1 and 2, respectively. The diagnosis of HSP was based on previously published criteria (Gasser *et al.*, 2010). Standardized clinical assessments were applied and included demographic characteristics, family history, pedigree, developmental history, and clinical symptoms. For a subset of patients, brain and spinal cord MRI were performed and the Spastic Paraplegia Rating Scale (SPRS) score was assessed (Schüle *et al.*, 2006). All participants have signed an informed consent form prior to enrollment, and the institutional review boards have approved the study protocols.

### Genetic and data analysis

DNA was extracted from peripheral blood using a standard salting-out procedure. In the HSP group, samples from 379 patients initially went through panel sequencing of HSP-associated genes. In total, 1,575 samples including 400 HSP and 1,175 control samples went through whole exome sequencing (WES). For exome target enrichment, SureSelect Human All Exon V4, V5, Nextera Rapid Capture Exomes and TruSeq Exome (Illumina) kits were used, and the sequencing was performed on Illumina HiSeq 2000/2500 platforms (San Diego, CA, USA). The sequence reads were aligned to the human reference genome (GRCh37/hg19) using Burrows-Wheeler Aligner (Li and Durbin, 2009). The calling of variants and annotation were done using Genome Analysis Toolkit and ANNOVAR, respectively (McKenna *et al.*, 2010; Wang *et al.*, 2010).

The initial selection of variants in the cohorts, was based on the identification of missense and protein truncating alleles in *SPG7* (OMIM # 602783, NM_003119.3) with minor allele frequency less than 0.01 in the gnomAD (Karczewski *et al.*, 2020). Using VarSome (Kopanos *et al.*, 2019), the variants have been classified according to the American College of Medical Genetics and Genomics guidelines. The variants classified as “Likely Benign” and “Benign” were excluded from further analysis. The intronic splicing variants with uncertain significance higher than ±3 were also excluded. Then, we aimed to examine whether carriers of *SPG7* variants may carry other variants in other HSP-associated genes or in genes linked to similar neurogenetic disorders which may involve spasticity. For this purpose, we searched for variants in 787 genes associated with such neurogenetic disorders (Supplementary Table 3) in HSP patients who carried at least one *SPG7* allele. Variant calls with less than 30x depth of coverage, a genotype quality of less than 97 and less than 25% genotyping frequency were excluded from the analysis. All the detected variants were visually inspected with the Integrated Genomics Viewer and suspicious variants were validated using Sanger Sequencing. To check for relatedness in the control group and in HSP families with complex inheritance of *SPG7*, we used *somalier* (Pedersen *et al.*, 2019). To test for the presence of population stratification within our cohort, we used PCA on filtered common variants from WES data. Genetic ancestry was determined by comparing it with HapMap (Nature, 2007).

Gene Ontology (GO) enrichment analysis was carried out using g:Profiler (Raudvere *et al.*, 2019), with Benjamini-Hochberg adjusted *p*-values for statistical significance set at < 0.05. For protein-protein interaction/network analysis, STRING (Szklarczyk *et al.*, 2019) (without the Textmining feature) and GeneMANIA (Warde-Farley *et al.*, 2010) were used. In order to predict the presence of important domains and sites of the corresponding protein, as well as multiple protein sequence alignment, we applied InterPro (Mitchell *et al.*, 2019) and Clustal Omega tools, respectively (Sievers *et al.*, 2011). A 3D atomic model of TBCE (a.a. 97-443) was built using the automated server I-TASSER (Yang *et al.*, 2015). The model was derived from the coordinates of different LRR domains sharing 12-20% identity. A second model of the TBCE LRR domain (a.a. 97-347) was also built with SWISS-MODEL (Waterhouse *et al.*, 2018) using the structure of the plant receptor BRI1 (pdb 3rj0, 33% sequence identity for this segment). The atomic coordinates of the TBC CAP-Gly domain (pdb 4b6m), TBCE Ubl domain (pdb 4icu) and human AFG3L2 (pdb 6nyy) were downloaded from the Protein Data Bank. The steric clashes induced by each mutation were evaluated using the “mutagenesis” toolbox in PyMol v. 2.3.5.

To study genotype-phenotype correlations in *SPG7*, we removed carriers of other pathogenic/likely pathogenic variants in HSP-related genes to avoid bias by other, non-SPG7 variants. We compared the following groups of patients: 1) carriers of homozygous variants, 2) carriers of compound heterozygous variants, and 3) carriers of heterozygous variants. Since it was previously suggested that HSP patients with the p.(Ala510Val) variant have a milder phenotype (Roxburgh *et al.*, 2013), we also compared carriers of this variant to other SPG7 patients. In addition, due to the report of differences in clinical presentations between SPG7 patients with loss-of-function (LoF) and missense variants, patients were classified and compared based on the variant type including patients with: 1) one missense, 2) one LoF, 3) one-missense and one-LoF 4), two missense, and 5) two LoF variants.

### Statistical analysis

For the analysis of binary variables, chi-square and Fisher’s Exact test were used, and for continuous variables Mann-Whittney U and Kruskal-Wallis tests were used, as required. SPSS was used to perform all statistical analyses. For the genotype-phenotype analysis of symptoms, Bonferroni correction for multiple comparisons was applied and corrected *p*-value threshold was set to < 0.0005.

## Results

### Identification of bi-allelic and monoallelic *SPG7* variant carriers with HSP

Out of 585 HSP patients, potentially pathogenic rare *SPG7* variants were identified through either panel sequencing or WES in 38 patients (6.5%) with homozygous or compound heterozygous variants and in 21 patients (3.6%) with heterozygous variants. In order to further compare frequencies of *SPG7* variants between patients and controls, we only included samples that went through WES, since controls did not go through panel sequencing, and since the panel sequencing did not always include *SPG7*. In index (unrelated) cases of HSP who underwent WES (n=291), 48 pathogenic/likely pathogenic *SPG7* alleles were detected compared to 10 in unrelated controls (out of n=580) who went through WES (OR=11.25, 95%CI= 5.60-22.62, *p*<0.0001). After excluding bi-allelic *SPG7* mutation carriers, we identified 15 patients (n=373, 4.02%) with heterozygous pathogenic/likely pathogenic *SPG7* variants vs. 15 (n=1,175, 1.28%) in controls (OR=3.24, 95% CI=1.56-6.69, *p*=0.0008, Table 1). When further analyzing only index cases, 13 (4.81%) heterozygous pathogenic/likely pathogenic *SPG7* variant carriers were identified in index cases with HSP who underwent WES (n=270) vs. 10 (1.72%) carriers which were identified out of 580 unrelated controls (OR=2.88, 95%CI=1.24-6.66, *p*=0.009), supporting a potential role for heterozygous pathogenic/likely pathogenic *SPG7* variants in HSP, or indicating potential digenic/oligogenic inheritance. Alternatively, it may also suggest an undetected *SPG7* variant such as deep intronic variant affecting splicing, copy number variants or missed coding variants. The heterozygous *SPG7* p.(Ala510Val) variant was found in 10 (3.70%) of the index cases vs. 5 (0.85%) in unrelated controls (OR=4.42, 95%CI=1.49-13.07, *p*=0.005). Supplementary Table 4 and Figure 1 details these variants in all cohorts.

**Table 1.**
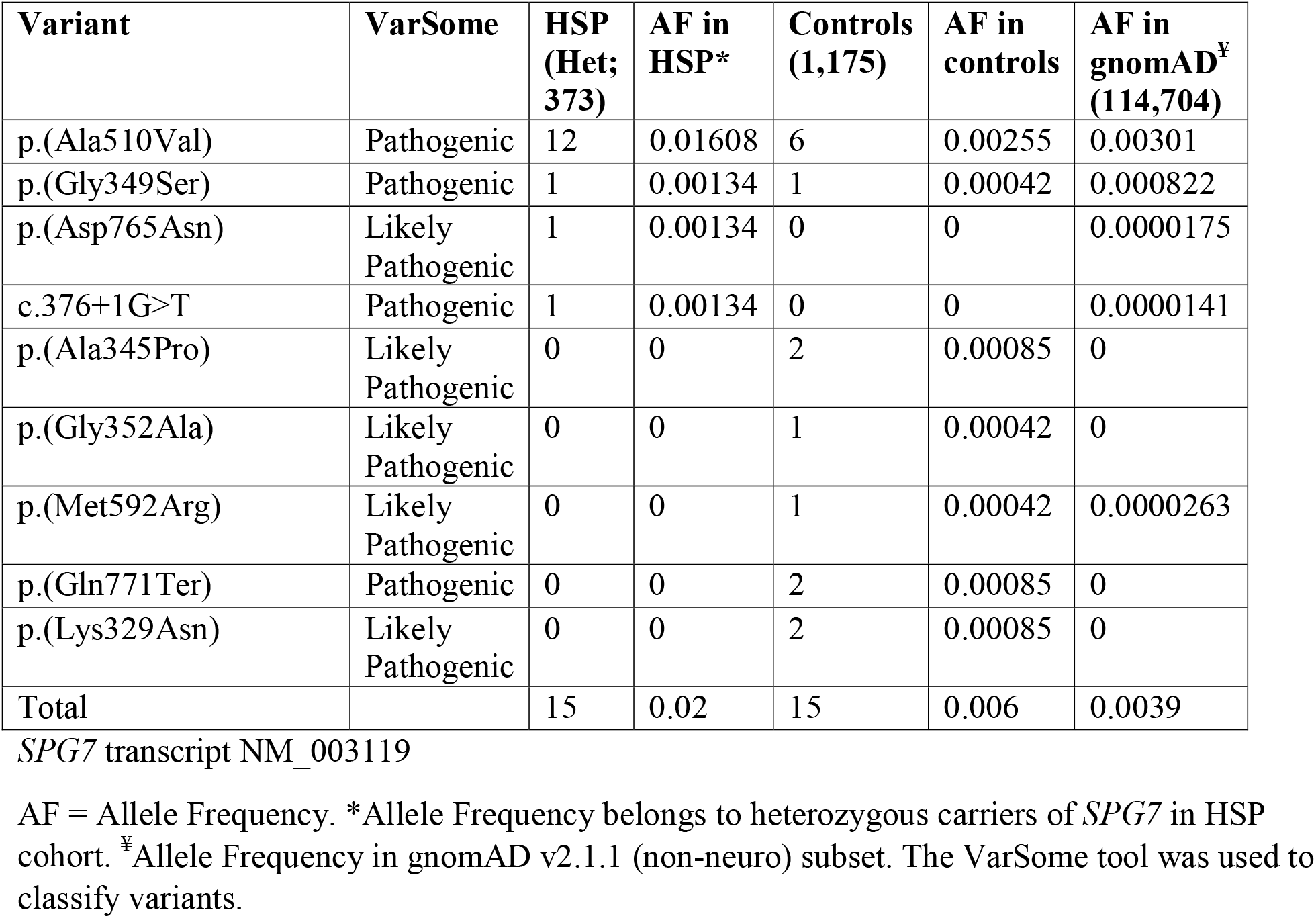
Heterozygous pathogenic and likely pathogenic *SPG7* variants identified in HSP patients and controls.

**Figure 1.**
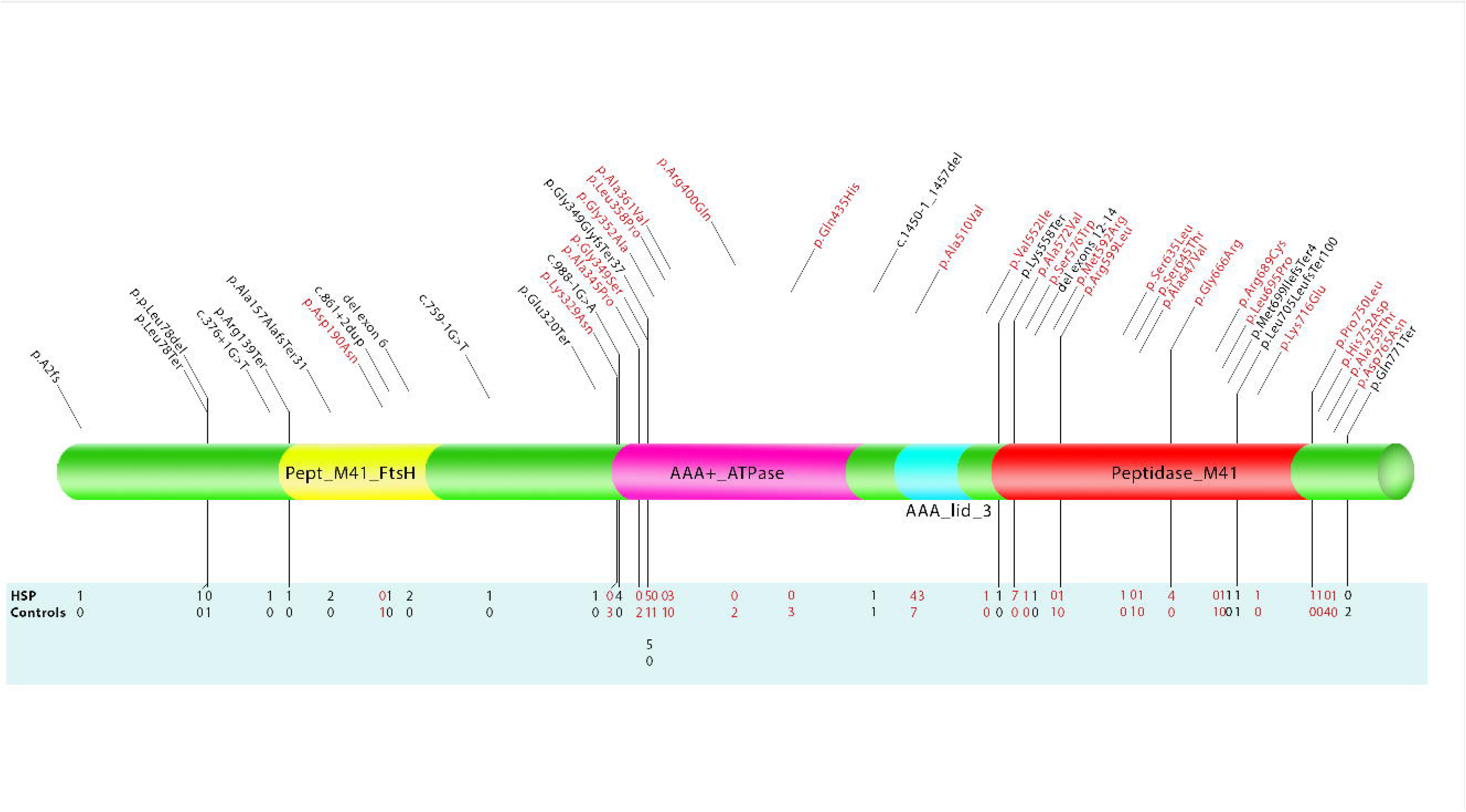
Schematic representation of the locations of *SPG7* variants in HSP patients and controls. InterPro predicted four domains for SPG7 including Pept_M41, AAA+_ATPase, AAA_lid_3 and Peptidase_M41 domains. Missense variants are indicated in red while loss of function variants in black. The below numbers represent the number of individuals carrying the specific variant.

### Evidence for potential digenic inheritance and modifier effects in SPG7-associated HSP

We further hypothesized that the overrepresentation of heterozygous *SPG7* variants in HSP patients vs. controls could be due to digenic inheritance, i.e. that patients with heterozygous *SPG7* variants may also carry variants in other HSP or similar neurogenetic disorders-associated genes (Supplementary Table 3). We found four index cases with heterozygous *SPG7* variant who also carried pathogenic variants in other genes that are associated with spastic paraplegia/ataxia (Table 2 details the patients’ symptoms and mutations), compared to zero in controls (OR=19.58, 95%CI=1.05-365.13, *p*=0.0031; Fisher’s Exact test after Haldane-Anscombe correction). One of these four families (01-013) had two affected members who carried a pathogenic variant in *BSCL2* (OMIM # 606158). One of the affected members of this family carried an *SPG7* p.(Ala510Val) variant while the other one was a non-carrier. In addition to the core symptoms, the *SPG7* variant carrier also presented with swallowing difficulties, upper extremity weakness, hyperreflexia and lower motor neuron features. One patient with heterozygous *SPG7* variant also carried two variants in *TBCE* (OMIM # 604934, Table 2), which is known to cause Encephalopathy (OMIM # 617207) with spasticity. One of the two *TBCE* variants was annotated as pathogenic, while the other was annotated as likely benign. This patient presented with clinical features that fit both *SPG7* and *TBCE* (Table 2). In addition, this patient presented with nephropathy and focal segmental glomerulosclerosis, which have not been previously reported in *SPG7* and are a very rare clinical finding in HSP (Efstratiadis *et al.*, 2006). We could not find a pathogenic variant in a gene related to nephropathy in this patient. Two other patients carried a *SPAST* mutation (Table 2). The average age at onset (AAO) of these four potentially digenic patients was 2.7 ± 2.1 y, compared to 28.0 ± 17.9 y in all other patients with *SPG7* variants (*p*=0.060, Mann-Whitney U test). Upper extremity weakness (2/2 vs 3/38; *p*=0.013) and hyperreflexia (3/3 vs 11/39; *p*=0.032) were more common among the potentially digenic patients with available data. None of these differences was statistically significant after correction for multiple comparisons.

**Table 2.** Genetic and clinical characteristics of HSP patients who carried at least one *SPG7* allele along with pathogenic non-SPG7 variant(s) that could explain the disease.

| Patients ' (in-house) ID | Symptoms | OMIM | Identified Gene Variant (Inheritance mode) | Novelty | VarSome | CADD | MutationTaster | Identified *SPG7* variants (Inheritance mode) | VarSome / Pathogenicity of *SPG7* variant in previous studies |
| --- | --- | --- | --- | --- | --- | --- | --- | --- | --- |
| FSP 01-013 | LEW, LES, LEH, Extensor Plantar Responses, Abnormal Bladder Function, Ankle Clonus, swallowing difficulties, upper extremity weakness, upper extremity hyperreflexia, amyotrophy or lower motor neuron features, Abnormal EMG results<br>AAO: 5 | LEW, LES, LEH, Hyperreflexia, Extensor plantar responses, Pes cavus, Decreased vibratory sense, Distal limb muscle weakness and atrophy, AAO: 8-40 | ***BSCL2***<br>NM_032667<br>c.269C>T;(p.[Ser90Leu])<br>(Het) | Reported (Cho *et al.*, 2007) | Pathogenic | 34 | 0.5876 (Disease Causing) | c.1529C>T;(p.[Ala510Val])<br>(Het) | Pathogenic / (Bonn *et al.*, 2010; Klebe *et al.*, 2012) |
| FSP 01-171 | LES, LEH, Abnormal Bladder Function, Ankle Clonus, upper extremity hyperreflexia, Mild vibratory loss<br>AAO: 2 | LEW, LES, LEH, Extensor Plantar Responses, Pyramidal signs, Cognitive decline, Decreased vibratory sense, Deficits in language expression, AAO: variable | ***SPAST***<br>NM_014946<br>c.1212_1216del; (p.[Asn405LysfsTer36])<br>(Het) | Novel | Pathogenic | - | - | c.1529C>T; (p.[Ala510Val])<br>(Het) | Pathogenic / (Bonn *et al.*, 2010; Klebe *et al.*, 2012) |
| FSP 01-208 | NA | - | *SPAST* deletion of exons 5-7 (Het) | - | - | - | - | 2293G>A; (p.[Asp765Asn]) (Het) | Likely Pathogenic / - |
| FSP 04-012 | LEW, LES, LEH, Extensor Plantar Responses, Ankle Clonus, Speech Delay or Abnormality, Learning Disability, progressive cognitive deficits, deafness, dysarthria, upper extremity weakness, upper extremity hyperreflexia, nephropathy, Focal segmental glomerulosclerosis SPRS: 37 AAO: 1 | Distal amyotrophy, Encephalopathy, Delayed psychomotor development, Motor regression, Spinal muscular atrophy, Intellectual disability, Ataxia, Spastic tetraplegia, Dysarthria, Absence of speech, Cerebellar atrophy, Thin corpus callosum, Axonal peripheral neuropathy, AAO: Infantile | *TBCE* NM_003193 c.661G>C; (p.[Val221Leu]) & c.1537C>T; (p.[Gln513Ter]) (Comp Het) | Novel & Novel | Likely Benign & Pathogenic | 15.44 & 41 | 0.5876 (Disease Causing) & 0.81 (Disease Causing) | c.1796G>T; (p.[Arg599Leu]) | Likely Pathogenic / - |

**LES** = Lower Extremity Spasticity; **LEH** = Lower Extremity Hyperreflexia; **LEW** = Lower Extremity Weakness; **SPRS** = Spastic Paraplegia Rating Scale; **Het** = Heterozygous. **Comp Het** = Compound Heterozygous ; **OMIM** = Online Mendelian Inheritance in Man; **AAO** = Age at Onset; **CADD** = Combined Annotation Dependent Depletion; **NA**: Not Available.

To examine whether *SPAST, BSCL2* and *TBCE* (the genes in which we found additional mutations in patients who also carry *SPG7* heterozygous mutation, Table 2), are involved in specific cellular pathways in which *SPG7* is also involved, we performed pathway enrichment analysis. We found enrichment mainly in axonal transport and cellular organization (adjusted *p*<0.05, Supplementary Table 5), with SPAST and SPG7 having the highest enrichment. Both SPAST and SPG7 play a major role in axonal transports and involved in biosynthesis, assembly and arrangement of macromolecules and cellular membrane and other components (Supplementary Table 5). Moreover, Spastin (SPG4) and Paraplegin (SPG7) contain the same ATPase domain where they participate in diverse cellular processes (IPR003959, IPR003593).

To identify additional patients who potentially have HSP due to digenic inheritance involving *SPG7*, we examined very rare variants (AF<0.001) in genes that produce proteins that interact with SPG7 (Supplementary Table 6). We found very rare heterozygous variants with uncertain significance in three genes *(CACNA1A, AFG3L2and MORC2)* in heterozygous carriers of *SPG7* (Table 3). Among them, AFG3L2 had the closest interaction with SPG7 and both share similar features, including protein structure and domains, interacting proteins, and biological functions and pathways (Figure 2). Except for one homozygous *AFG3L2* variant (NM_006796), c.122G>A;(p.[Arg41Gln]) with gnomAD AF 0.00000481 (predicted as polymorphism by MutationTaster), which was identified in one control (who did not carry an *SPG7* variant) out of 1,175, no rare variant was detected in *CACNA1A, AFG3L2* and *MORC2* genes in our controls. Yet, to confirm their potential pathogenicity in digenic HSP, additional genetic and functional studies are needed.

**Table 3.** Variants in *SPG7* interacting genes which were identified in HSP patients with heterozygous *SPG7* variants.

| Patients' (in-house) ID | Gene (known phenotype) | Variant | gnomAD (non-neuro) AF | CADD | MutationTaster | SIFT | GERP | SPG7 variant | Evidence of interaction with SPG7 |
| --- | --- | --- | --- | --- | --- | --- | --- | --- | --- |
| FSP-04-053 | *AFG3L2*<br>(Spinocerebellar ataxia 28 # 610246) | NM_006796<br>c.2114T>C;(p.[Ile705Thr]) * | 0 | 29.1 | 0.81 (Disease causing) | 0.006 (Damaging) | 5.6 | c.376+1G>T | (Sacco *et al.*, 2010; Magri *et al.*, 2018; Patron *et al.*, 2018) |
| FSP-01-190 | *CACNA1A*<br>(Spinocerebellar ataxia 6 # 183086) | NM_001127221<br>c.4981C>T;(p.[Arg1661Cys]) | 0 | 25.3 | 0.81 (Disease causing) | 0 (Damaging) | 4.9 | p.(Ala510Val) | (Coutelier *et al.*, 2017) |
| FSP-04-045 | *MORC2* (Charcot-Marie-Tooth disease, axonal, type 2Z # 616688) | NM_014941<br>c.2716C>T;(p.[Arg906Cys]) | 9.61e-6 | 23.7 | 0.5876 (Disease causing) | 0.19 (Tolerated) | 4.3 | p.(Lys716Glu) | GeneMANIA |

\*The variant was reported from one hereditary ataxia patient (Iqbal *et al.*, 2017).
**AF** = Allele Frequency; **SIFT** = Sorting Intolerant From Tolerant; **GERP** = Genomic Evolutionary Rate Profiling; **CADD** = Combined Annotation Dependent Depletion;

**Figure 2.**
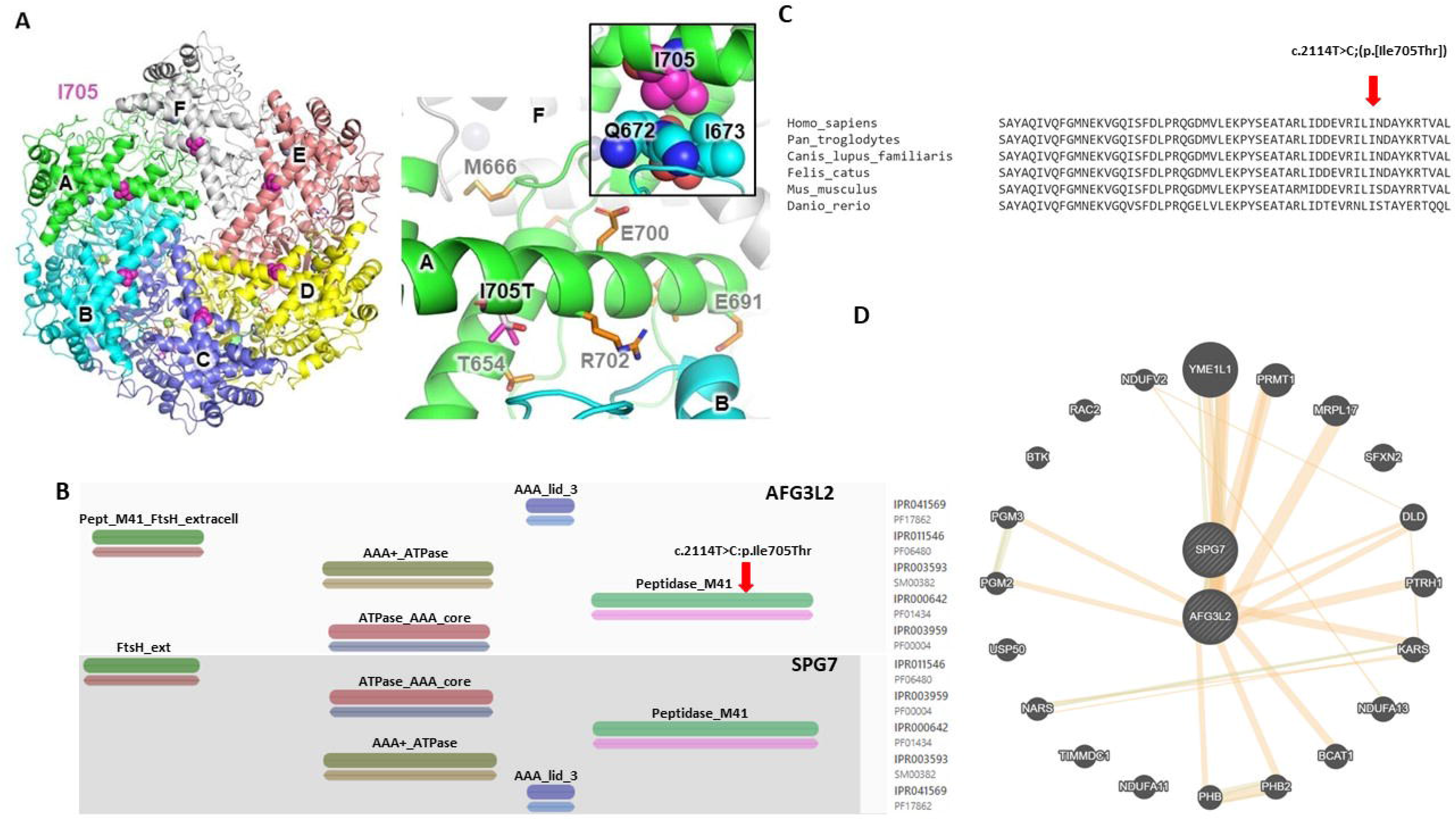
The mutation p.Ile705Thr in *AFG3L2*. **A**. Cryo-electron microscopy (cryo-EM) structure of human AFG3L2 (pdb 6nyy). The protein forms a homo-hexamer with the six chains labeled α-f. Ile705 is show in magenta. **B**. Close-up view of the p.Ile705Thr mutation site. The side-chain of Ile705 in chain A makes VdW contacts with a loop in chain B (small inset, top right). The mutation (white) introduces no clash, but makes the interface less hydrophobic. SCA28 mutation sites are shown in orange. Cartoon images produced using PyMOL v.2.3.5. **C**. Domain prediction by InterPro revealed that SPG7 and AFG3L2 share similar protein domains. The variant in AFG3L2 occurred in Peptidase M41 (IPR000642) which belongs to metallopeptidase families. **D**. Sequence alignment of AFG3L2 orthologs, showing conservation of Ile705. The AFG3L2 mutation occurred in an amino acid that is highly conserved among species. **E**. Network construction from *SPG7* using GeneMANIA showed that AFG3L2 and SPG7 are partner proteins that interact with other genes in multiple pathways.

### Structural analysis of *TBCE* and *AFG3L2* variants

Among the non-HSP genes that may contribute to oligogenic inheritance or epistasis, *AFG3L2* and *TBCE* were the strongest candidates. To further examine whether the two *TBCE* variants identified in family 04-012 (Table 2) may be pathogenic we performed structural analysis of their potential effects on TBCE structure and function. The human tubulin cofactor E (TBCE) consists of a N-terminal CAP-Gly domain, followed by a leucine-rich repeat (LRR) domain and a C-terminal ubiquitin-like (Ubl) domain (Figure 3A). The structures of TBCE alone and in complex with α-tubulin and TBCB were determined at low resolution by electron microscopy (Serna *et al.*, 2015). The complex structure shows that α-tubulin binds to the CAP-Gly domain as well as the concave surface of the LRR domain. This interaction enables the TBCE-TBCB complex to dissociate the α/β tubulin heterodimers into monomers that can be degraded by the ubiquitin-proteasome system (Mi *et al.*, 2009). To investigate the impact of the potential HSP-related variant p.(Val221Leu) on TBCE’s structure and function, we performed *in silico* mutagenesis of the residue in two homology models of TBCE. Val221 is located on the “exterior” side of the LRR domain, opposite to the side of interaction with tubulin (Figure 3A). In the homology model generated with I-TASSER, the side-chain of Val221 points towards the solvent. The variant Val to Leu results in a clash with a helix in an adjacent repeat (Figure 3B). In the homology model generated with SWISS-MODEL using the homologous LRR domain of the plant steroid hormone receptor BRI1 (pdb 3rj0, 33% sequence identity), Val221 is located in the hydrophobic core, and the p.(Val221Leu) variant also leads to steric clashes. In both models, our prediction is that the variant destabilizes the domain and might inactivate it. The mutation p.(Gln513Ter) would result in a 15 amino acid deletion in the Ubl domain of TBCE (Figure 3A). The crystal structure of the Ubl (pdb 4icu) shows that this segment comprises a helix and the C-terminal β strand, which is typically involved in protein-protein interactions (Trempe, 2011). Deletion of this segment would therefore completely unfold the Ubl and disrupt its function. The Ubl domain might be involved in shuttling tubulin to the proteasome, given that the proteasome subunit Rpn10 binds to ubiquitin and Ubl domains (Riedinger *et al.*, 2010; Chen *et al.*, 2019). However, the Ubl domain of TBCE has only 21% sequence identity with ubiquitin, and comparison of the TBCE Ubl with the complex of Rpn10 and the Ubl of UBQLN2 (14% sequence identity) reveals that most residues interacting with Rpn10 are not conserved between UBQLN2 and TBCE. Therefore, the TBCE Ubl is unlikely to bind to the proteasome. Its function thus remains unknown.

**Figure 3.**
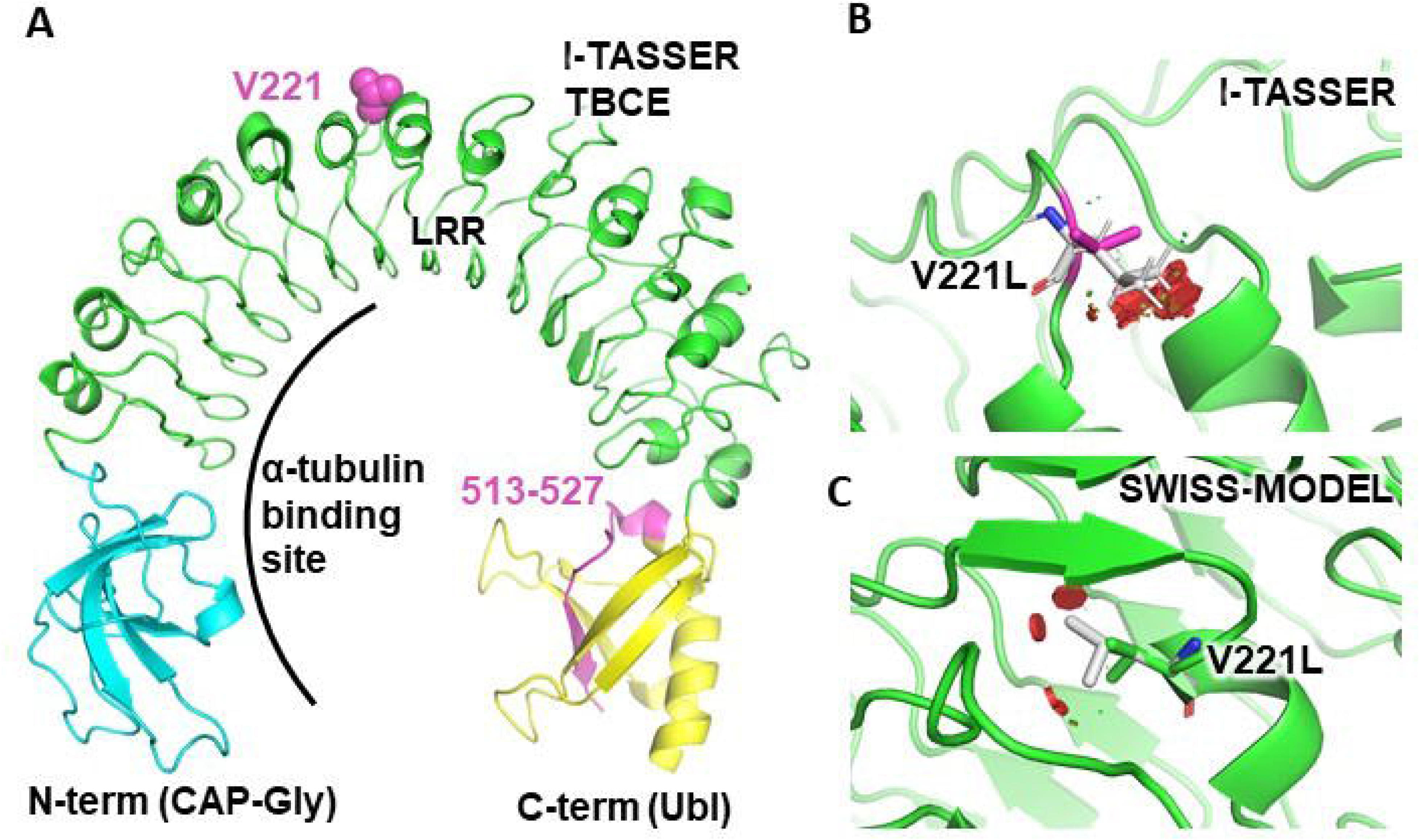
Structural analysis of the identified mutations in TBCE. **A**. Homology model of TBCE a.a. 90-443 (green) produced using I-TASSER and merged with the structure of a CAP-Gly domain (a.a. 1-90, cyan, pdb 4b6m) and crystal structure of the C-terminal Ubl domain (a.a. 444-527, yellow, pdb 4icu). Mutation sites are in magenta. Val221 is in the leucine-rich repeat (LRR) domain. The p.Gln513Ter mutation removes a.a. 513-527, which form part of a helix and the C-terminal β strand. The α-tubulin binding site derived from a low-resolution EM structure is shown. **B**. Close-up view of the p.Val221Leu mutation site in the I-TASSER model. The mutated residue is shown in white. The mutation would create clashes (red disks) with a helix in an adjacent repeat. **C**. Close-up view of the p.Val221Leu mutation site in the SWISS-MODEL homology model. Here the side-chain of Val221 points in the hydrophobic core, and the mutation results in clashes as well. Cartoon images were produced using PyMOL v.2.3.5.

AFG3L2 is a subunit of the m-AAA protease complex, which cleaves proteins in the mitochondrial inner membrane (Koppen *et al.*, 2007). It consists of a N-terminal segment that anchors them to the membrane, followed by an ATPase and protease domains that assemble into a hexamer. The structure of the soluble domains of human AFG3L2 bound to a substrate was determined by cryo-electron microscopy (cryo-EM) at high-resolution and revealed how its protease and ATPase domains coordinate to pull in substrates for proteolysis (Puchades *et al.*, 2019). The mutation p.(Ile705Thr) locates to a helix in the central protrusion of the protease domain at the interface of the protease in another subunit (Figure 2A). The side-chain of Ile705 forms hydrophobic interactions with a loop formed by Gln672 and Ile673 (Figure 2B). The mutation p.(Ile705Thr) results in no steric clash, but would make the interface less hydrophobic. Intriguingly, Ile705 is located in a “hotspot” of SCA28-associated mutations such as p.(Thr654Ile), p.(Met666Arg/Val/Thr), p.(Pro688Thr), p.(Tyr689His/Asn), p.(Glu691Lys), p.(Glu700Lys), and p.(Arg702Gln) (Figure 2B). All of these mutations strongly reduce the ATPase and degradation rates of AFG3L2, and the mutations p.(Met666Arg), p.(Pro688Thr), and p.(Glu691Lys) prevent formation of hexamers (Puchades *et al.*, 2019). Thus, the mutation p.(Ile705Thr) may also affect the stability of the AFG3L2 hexamer and may compromise the functional coupling between the protease subunits.

### Genotype-phenotype correlations among *SPG7* mutation carriers

Next, we examined whether genotype-phenotype correlations exist between different groups of *SPG7* variant carriers. Clinical data was available for 12 heterozygous carriers, 9 homozygous carriers (7 of whom are homozygous carriers of the p.(Ala510Val) variant), and 22 compound heterozygous carriers. Table 4 details the clinical data for each of these groups, each time comparing one group to the other two. After correction for multiple comparisons, there were no statistically significant differences between the groups. However, some differences between heterozygous carriers and bi-allelic variant carriers were notable. While upper extremity ataxia (37.9%) and intent tremor (30%) were relatively common in bi-allelic carriers of *SPG7* variants, none of the heterozygous carriers showed these abnormalities. On the other hand, two heterozygous carriers presented with motor developmental delay which was not found in any bi-allelic patients. Moreover, patients with heterozygous *SPG7* variants had younger AAO compared to bi-allelic patients (16.5 vs. 33.8 y, *p*=0.021). Cerebellar atrophy was the most common imaging finding among bi-allelic patients (22.7%).

**Table 4.**
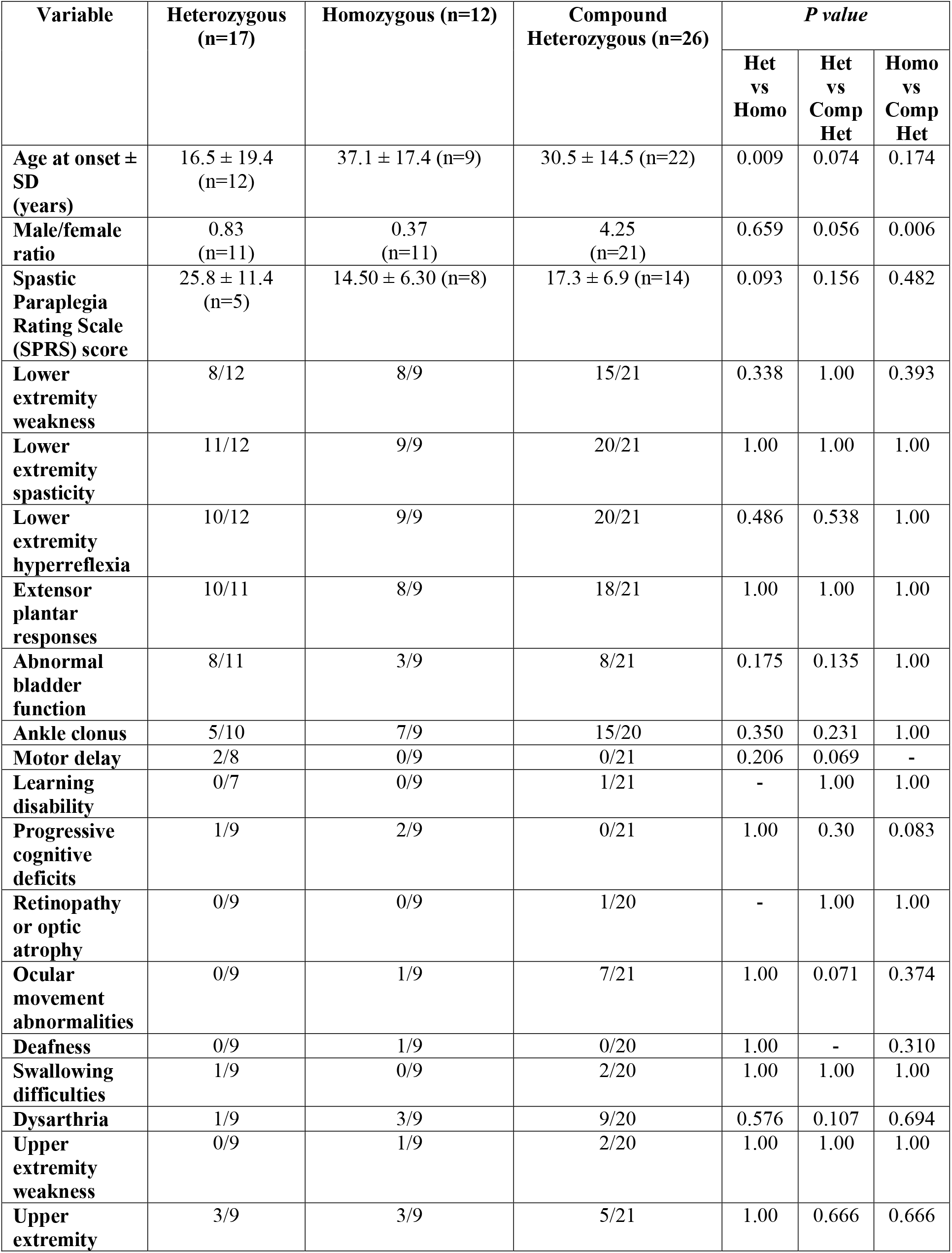
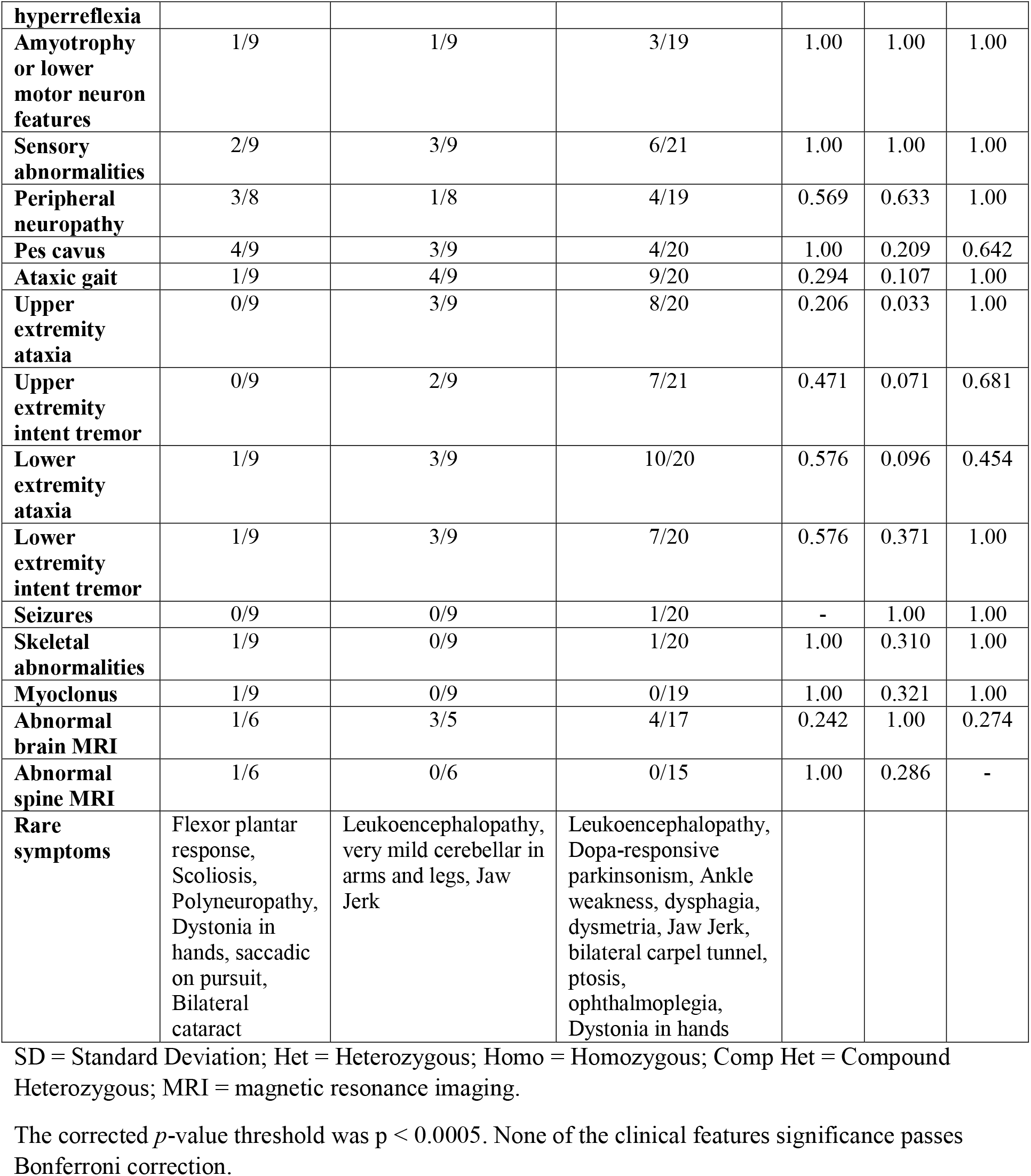
Genotype-phenotype correlation analysis in HSP patients according to *SPG7* variants
status.

We further examined whether there are differences between subgroup of patients, based on the type of variants that they carried (missense, loss-of-function) and the presence of the most common variant, p.(Ala510Val) which was carried by 34 (53.9%) of patients with at least one allele. No statistically significant differences were identified after correction for multiple comparisons (Table 4, Supplementary Table 8), likely due to the small number of patients in each of these groups.

## Discussion

The current study summarizes genetic and clinical data on *SPG7* from a large Canadian cohort of 585 patients, of which 6.5% carried bi-allelic *SPG7* variants. Our findings show that the number of pathogenic/likely pathogenic *SPG7* alleles in HSP patients is higher than in controls, even when considering only heterozygous carriers in index HSP patients vs. unrelated controls, suggesting potential dominant or digenic inheritance in some cases. Along these lines, we found that some of the heterozygous carriers of *SPG7* pathogenic variants with HSP also carried other potentially pathogenic variants in other genes, a phenomenon which was not observed in controls. These findings, which require replication, may suggest digenic inheritance in HSP associated with *SPG7*. Of note, we cannot rule out that in some of the heterozygous carriers of *SPG7* variants, an additional *SPG7* variant exists that was not detected through WES due to the limitation of WES to detect deep intronic and large structural variants.

The four patients with heterozygous *SPG7* variants who also carried other variants in genes related to HSP phenotype had younger AAO (2.7 vs 28 y) and higher average SPRS score (37 vs 18.1), indicating that their disease is more severe than those who did not carry variants in two genes. Upper extremity weakness and hyperreflexia are relatively common in these potentially digenic patients compared to other carriers of mono-allelic and bi-allelic *SPG7* alleles. We further examined the possibility of genetic interaction/modification by performing biological pathway enrichment analysis. SPAST and SPG7, whose variants co-occurred in three HSP patients, share a similar ATPase domain and closely interact in multiple pathways. In the control cohort, none of the participants carried pathogenic variants in the genes which were identified in the HSP patients *(SPAST, BSCL2*, and *TBCE)*. Overall, these findings may suggest either digenic inheritance, or epistasis between heterozygous *SPG7* variants and other HSP-related genes, as previously reported in SPG4 (Svenson *et al.*, 2004; Erichsen *et al.*, 2007; Newton *et al.*, 2018). Digenic inheritance has been recently suggested in neurodegenerative disorders including spinocerebellar ataxia and Charcot-Marie-Tooth (CMT), when the diseases were initially considered as a simple Mendelian monogenic disorder (Figueroa *et al.*, 2017; Nam *et al.*, 2018). More recently, it was demonstrated that the *EXOC4* gene may be involved in complex inheritance in axonopathies, including HSP and CMT (Bis-Brewer *et al.*, 2020).

By combining interaction and genetic analyses, we have identified heterozygous carriers of *SPG7* who also carried variants in genes potentially interacting with SPG7, including *AFG3L2, CACNA1A, PNMA1* and *MORC2*. It is possible that the co-occurrence of the heterozygous variants of the above-mentioned genes with *SPG7* heterozygous variants may lead to HSP. These results require replications in additional cohorts, as well as additional functional evidence. The pathway enrichment, domain prediction, protein network and protein conservational analysis showed that AFG3L2 is the strongest potential candidate interacting with SPG7. This finding is further supported by functional studies, demonstrating the potential interaction of these two proteins within the mitochondria (Martinelli *et al.*, 2009; Patron *et al.*, 2018). SPG7 and AFG3L2 exert overlapping substrate specificities, hence the expression level of AFG3L2 and SPG7 might be important in cell-type specificity in disorder. Dominant *AFG3L2* mutations cause spinocerebellar ataxia type 28 (SCA28; OMIM # 610246) whereas biallelic mutation may affect the interaction of SPG7 and AFG3L2 and cause spastic ataxia 5 (SPAX; OMIM # 614487), a disease whose phenotype includes features of both SCA28 and SPG7. A recent study reported a patient with heterozygous variants in both genes with syndromic parkinsonism and optic atrophy (Magri *et al.*, 2018). SPG7 and AFG3L2 are components of mitochondrial m-AAA proteases and they can assemble hetero-oligomeric proteolytic complexes with SPG7 (Patron *et al.*, 2018). Together with the findings in the current study, these data suggest that *SPG7-AFG3L2* digenic variants may be a cause of HSP and similar disorders, and that individuals with heterozygous *SPG7* variants with neurodegeneration should be specifically screened for *AFG3L2* variants.

In this study we also report several rare clinical features of SPG7, including dopa-responsive parkinsonism, dysphagia, dysmetria, jaw jerk, ptosis, ophthalmoplegia, optic atrophy and hands dystonia in bi-allelic *SPG7* patients. A few studies previously reported some of these symptoms in patients with *SPG7* mutations (Van Gassen *et al.*, 2012; Pfeffer *et al.*, 2014; Pedroso *et al.*, 2018; De la Casa-Fages *et al.*, 2019; Jacinto-Scudeiro *et al.*, 2019). Optic atrophy and dopa-responsive parkinsonism were also reported in a patient with concurrent *AFG3L2* and *SPG7*heterozygous variants (Magri *et al.*, 2018). Similar to other genes involved in HSP, *SPG7* may be involved in a continuum of spastic neurogenetic disorders, and *SPG7* variants should not be ruled out only because of the presence of rare clinical manifestations.

Our study has several limitations. Despite being one of the world’s largest cohorts of HSP, the total number of SPG7 patients is still relatively small, especially for genotype-phenotype studies. In addition, not all our HSP cohort went through WES, which prevented the participation of all patients in some of the analyses. One limitation of WES is that it cannot properly detect genetic variants such as large copy number variants (CNVs). While we did analyze the data with ExomeDepth (Plagnol *et al.*, 2012), a computational tool for CNV detection in WES data, and did not identify CNVs, we cannot rule out that some of our supposedly heterozygous carriers of *SPG7* variants carry an additional undetected *SPG7* variant such as a large deletion.

To conclude, our results suggest that the inheritance of *SPG7* may be complex, and include dominant or digenic inheritance, on top of the known recessive inheritance. One of the most intriguing findings, which requires additional replications, is the potential digenic inheritance with *AFG3L2* variants. The relatively high allele frequency of some of the pathogenic *SPG7* variants in the general population, the results of the genotype-phenotype correlation analysis, and a recent functional study (Magri *et al.*, 2018) support this possibility. Future studies will benefit from whole genome sequencing, which will allow for identifying CNVs, and deep intronic variants that can lead to aberrant splicing, as well as for comprehensive investigation of complex inheritance in SPG7 and other forms of HSP.

## Data Availability

The full protocol is available upon request.

## Acknowledgements

We thank the patients and their families for participating in this study. MAE and FA are funded by the Fonds de Recherche du Québec-Santé (FRQS). KMB holds a Canada Research Chair (Tier 1) in Rare Disease Precision Health. JFT holds a Canada Research Chair (Tier 2) in Structural Pharmacology. GAR holds a Canada Research Chair (Tier 1) in Genetics of the Nervous System and the Wilder Pen field Chair in Neurosciences. ZGO is supported by the Fonds de recherche du Québec-Santé Chercheur-Boursier award and is a Parkinson Canada New Investigator awardee. We thank D. Rochefort, H. Catoire, and V. Zaharieva for their assistance.

## Supplementary Tables and Figures

Supplementary Figure 1. The principal component analysis (PCA) on common variants was used to test for the presence of population stratification.

Supplementary Table 1. The genetic and clinical data of HSP subjects were included in this study.

Supplementary Table 2. The characteristics of control individuals.

Supplementary Table 3. The complete list of genes associated with spastic paraplegia phenotype. The list includes known HSP genes (https://omim.org/phenotypicSeries/PS303350), the list of in-house genes, gene panel for movement disorders (https://www.radboudumc.nl/en/patientenzorg/onderzoeken/exome-sequencing-diagnostics/exomepanelspreviousversions/exoompanels-huidige-en-voorgaande-versies/movement-disorders) and genes from HSPome (Novarino *et al.*, 2014).

Supplementary Table 4. The list of *SPG7* variants which were detected in all the subjects included in this study.

Supplementary Table 5. Gene Ontology (GO) term enrichment with respect to Biological Process using g:Profiler. *p* values were corrected for multiple testing using the Benjamini-Hochberg false discovery rate. Adjusted *p* values show the significance of enrichment with the threshold of *p*<0.05. The data source from Gene Ontology with Biological Process (BP) subontology was used to describe in which biological process the gene product participates.

Supplementary Table 6. The list of SPG7-interacting genes.

Supplementary Table 7. Gene Ontology (GO) term enrichment using g:Profiler (Benjamini-Hochberg adjusted *p*-values set at < 0.05).

Supplementary Table 8 **a**. The comparison of clinical characteristics in different groups based on variant type. **b**. The comparison of clinical characteristics in different groups based on p.(Ala510Val) variant.

